# Epistemic Trust, Mistrust and Credulity Questionnaire (ETMCQ) validation in French language: Investigating association with loneliness

**DOI:** 10.1101/2024.05.06.24306924

**Authors:** Christian Greiner, Vincent Besch, Marissa Bouchard-Boivin, Catherine Le Hénaff, Cécilia Von Rohr-De Pree, Nader Perroud, Paco Prada, Martin Debbané

**Author notes:** Corresponding author (CG). These authors contributed equally to this work (Christian Greiner, Vincent Besch). These authors also contributed equally to this work (Paco Prada, Martin Debbané).

## Abstract

The concept of epistemic trust is gaining traction in the mental health field. It is thought to play a foundational role as a resilience factor against the development and maintenance of psychopathology by facilitating social learnings useful to navigate in the modern world. The primary aim of this study is to validate in French language the Epistemic Trust, Mistrust, and Credulity Questionnaire (ETMCQ). We further investigate associations with key developmental and psychological factors (childhood trauma, mentalizing and attachment), besides possible mediating roles between childhood traumatic experiences and psychopathology and between loneliness and psychopathology. 302 participants were recruited for analysis via the online survey platform Prolific. Along with ETMCQ, measures of CTQ-SF, RFQ-8, ECR-R, UCLA-LS and SCL-90-R were administered. Confirmatory Factor Analysis and General Linear Model of Mediation were conducted. Our study shows that the ETMCQ represents a valid instrument to assess epistemic trust. We find an adequate replication of the original three-factor solution in a francophone population with a 12-item version, exhibiting satisfactory psychometric properties and external validity. We replicate previous findings that demonstrated epistemic trust’s attachment style related differences, as well as the mediating effect between childhood traumatic experiences and psychopathology. We also observed that epistemic trust mediates the well-described association between loneliness and psychopathology. We add momentum to the framework that considers epistemic trust as key underlying contributor to the maintenance or alleviation of psychopathology. Future research should investigate the ETMCQ in clinical population, where psychopathological expressions are severe, enduring and connected, and where identifying potential intercessors could help target and improve interventions.

## Introduction

In the past decade, the field of mental health appears to be increasingly focusing on social-cognitive abilities enabling individuals to form, maintain, lean on, and learn from interpersonal relationships and social networks [1]. The work of Fonagy and colleagues on the relationship between attachment, mentalizing and trust propose to re-examine the links between early adversity and the expression of psychopathology over the lifespan [2–5]. Building on a broad range of disciplines such as evolutionary anthropology [6], philosophy [7,8] and epistemology [9], but also on developmental psychology [10] and attachment research [11,12], Fonagy and colleagues propose an integrative theoretical model in which early adversity in attachment relationships, impaired mentalizing and psychopathology are developmentally interwoven through difficulties in attributing trust to a key sources of social information, that is, to attribute epistemic trust. This misattribution ultimately limits social learning and consequently interferes with adaptation and resilience. They define epistemic trust as “the capacity of the individual to consider knowledge conveyed by others as significant, relevant to the self, and generalizable to other contexts” [13]. They hypothesize that early adversity impacts the development of social communicative abilities, conferring patterns of mistrust and/or credulity, first to attachment figures [14], and then to other significant people in the individual’s social environment, preventing the individual to learn to adaptively navigate and learn from different social contexts [15,16]. The concept of epistemic trust does not just concern an open stance to trust any given information or general trust, but rather a more complex process that depends on the individual’s ability to consider the reliability of the source of information, the information’s relevance to the addresses, and the quality of the information in a calibrated fashion [16].

In order to empirically examine the epistemic trust theoretical framework, Campbell et al. [13] operationalize the concept through three possibly linked dimensions embodied within social communication: Epistemic Trust (ET), Epistemic Mistrust (EM) and Epistemic Credulity (EC). ET is characterized by selective and balanced receptivity to social learning opportunities within relationships, and the capacity to maintain confidence in the reliability and value of information from others. EM is defined as perceiving a source of information as untrustworthy, leading to individuals’ tendency to remain impermeable to the influence of others’ communication. EC entails decreased vigilance and discrimination vis-a-vis social signals, making the individual prone to misinformation and exploitation [17]. EM and EC are often sustained by patterns of inefficient mentalizing characterized by all or nothing views on mental states of self and others (psychic equivalence), concrete thinking (teleological mode) or decoupling between emotional needs and emotion expression (pretend mode). Campbell et al. [13] introduced the first self-report scale (15 items) to assess epistemic trust, the Epistemic Trust, Mistrust and Credulity Questionnaire (ETMCQ). Their results offer evidence for a three-factor structure, empirically substantiating the hypothesis of three distinct epistemic stances. The questionnaire yields good reliability and validity indices, and significant associations between both EM and EC and higher levels of childhood adversity, low mentalizing abilities, as well as insecure attachment and expressions of psychopathology. Post-hoc analysis identified that different attachment styles were associated with differences in epistemic stances. Both EM and EC partially mediated the link between early adversity and expression of psychopathology. To our knowledge, two ETMCQ translation and replication studies were conducted to date. Liotti et al. [18] translated and validated the scale into Italian, and Asgarizadeh et al. [19] into Persian. Both provide evidence for the instrument’s cross-cultural applicability, by reporting a three-factor structure similar to the model proposed in the original validation with slight differences on item loadings and total item number. Other studies which employ the ETMCQ with convergent psychological variables in large and representative populations or clinical samples convey further evidence supporting key assumptions regarding the relations of dysfunctional epistemic stances with adverse childhood experiences, insecure attachment, poor mentalization and psychopathology [17,20–23,18,24–27].

Within this framework, epistemic trust is thought to play a foundational role as a resilience factor against the development and maintenance of psychopathology, through its salutogenic effect in sustaining adaptive social learning. It could accordingly act as a proxy for a general psychopathology factor [4], or as a mediating “psychomarker” predicting outcomes of psychosocial interventions, not limited to psychotherapeutic treatment but to any intervention that depends on trust in others [28]. Currently, the interplay between attachment, childhood traumatic experiences and later outcomes remain difficult to grasp [29,30]. Whilst most clinical researchers and practitioners are familiar with attachment-related differences, the exact mechanisms linking to developmental psychopathology remain enigmatic [31–33]. If attachment-related differences reflect broader differences in establishing trust in communication - as John Bowlby had intuited [35] - rather than exclusively strategies for managing threats, their association with lifespan evolutions could become more intelligible [35–37]. There is strong evidence of an association between childhood trauma and unfavorable later-life consequences [38–40], where epistemic trust appears to significantly mediate the effect between early adversity and mentalizing abilities [17] as well as with psychopathology [13]. Other results are however mixed [41] and require further studies to inform the complex paths between early disruptive experiences and mental health difficulties. Research on well-being, both psychological and social, is gaining traction as a body of scientific literature putting emphasis on “living well” as opposed to the more conventional health-centered view of “getting rid of illness” [42]. It joins a broader movement encompassing recovery and salutogenesis, a path also thread by the mentalization research field [4,43]. Loneliness, a concept with a lengthy research record, is consequently reappraised as a key social factor playing against well-being, in a transdiagnostic fashion [44]. Described as “gnawing chronic disease without redeeming features” [45], it emerges as a serious global health concern, as it is a risk factor for a myriad of both physical and mental health conditions, rivaling well-established morbidity risk factors such as physical inactivity, smoking and obesity [46]. Interestingly, loneliness’ chief model of understanding advanced by Cacioppo and colleagues shows thought-provoking similarity with the epistemic trust construct, particularly “epistemic petrification”, an expression seeking to translate how the traumatic valence of early adversity resides in the solitude it entails when facing threat, when early adversity tends to convey the lesson that vigilance and impermeability to others’ influence may be the most adaptive way to navigate a dangerous social world [3]. In a similar vein, but from a cognitive behavioral standpoint, Cacioppo et al. theorize a self-reinforcing loop which would sustain the formation and maintenance of chronic loneliness [47]. They propose that loneliness can increase hypervigilance and cognitive biases towards social threat, leading lonely individuals to anticipate negative social interactions and remember more negative social information. As a result, lonely individuals may exhibit hostile or pessimistic behaviors which elicit exactly the unwanted responses from others that confirm their negative expectations. This loop has short-term self-protective features but over the long-term, overall consequence is that a negative self-image is established, along with a desire to avoid social contact, resulting in chronic feelings of loneliness [48]. Thus, besides being well suited for studying links between attachment, adversity and later psychopathology, the epistemic trust framework appears to lend itself very well to the inquiry of loneliness. Specifically, disentangling the question of causality and reciprocal influences [49,50] in the already firmly established link between loneliness and psychopathology [51] could be further investigated if were to hypothesize that epistemic trust stances play a mediating partition in these complex developmental transactions.

### Aims and Hypotheses

The first objective of the present study is to validate the ETMCQ in French language to provide a valid tool to francophone clinical and research settings. We expect to confirm a three-factor structure, referring respectively to the degree of trust, mistrust and credulity. Furthermore, we suppose that this three-factor structure will have satisfactory internal consistency and be invariant across time (1-year). We also anticipate that the ETMCQ scores converge with key developmental variables (levels of childhood traumatic experiences, attachment styles) and psychological processes (mentalization abilities), as theoretically proposed and empirically tested in previous studies.

Second, we seek to replicate Campbell et al. [13] main findings, expecting to observe that the ETMCQ subscales differ according to distinctive attachment dimensions and styles. We further expect the ETMCQ subscales to significantly mediate the association between childhood traumatic experiences and psychopathology.

Third, in line with the theory put forward by Fonagy and Allison [2], we investigate the potential association between epistemic trust and loneliness. We expect to find that the ETMCQ subscales significantly mediates the association between loneliness and the individual expression of psychopathology.

## Methods

### Participants

In this study, 372 participants were recruited via the online survey platform Prolific (https://www.prolific.com/). The inclusion criteria were to be 18 years of age or older, to be fluent in French, and to have never been hospitalized for psychiatric reasons. Participants received a financial compensation of 6.00 CHF for each test session, with a +25% bonus for the 1-year retest. In order to ensure the quality of the data they provided, several additional selection criteria were applied. First, the need for participants to have satisfactorily submitted at least 15 online surveys beforehand to ensure a good level of use of the online platform and reduce data entry errors. Then, nonsensical items were randomly inserted in the survey and response times were monitored in order to check for participants’ linguistic skills and attention. 70 participants failed these controls and were rejected. The final sample used for analyses included N=302 participants. All participants filled-in socio-demographic data and completed the initial set of 15 items of the ETMCQ and additional validity questionnaires. Recruitment period started April 21^st^, 2022, and ended January 19^th^, 2024. The study was approved by the Swiss Ethics Commission in Geneva under Project-id 2021-01100. Participants were submitted a written consent form approved by the Commission. According to its requirements, participants gave written consent on their involvement in the study and the use of their data for analysis and publication.

### Additional Measures

<u>Childhood Trauma Questionnaire - Short Form (CTQ-SF)</u> [52]: a 28-item self-report instrument validated for clinical and non-clinical populations. The questionnaire evaluates the presence and severity of five types of childhood traumatic experiences: Physical Abuse, Emotional Abuse, Sexual Abuse, Emotional Neglect and Physical Neglect. Individuals are asked on a 5-point Likert scale to indicate how often they suffered specific traumatic childhood experiences. The French validation of the instrument demonstrated that the five types of traumatic childhood experiences are valid and usable with French-speaking populations [53].

<u>Reflective Function Questionnaire (RFQ-8)</u> [54]: an 8-item self-report instrument to evaluate mentalization abilities by the degree of certainty and uncertainty with which individuals utilize mental state information to understand their own and other’s behaviour. It asks subjects to express their agreement with each item on a 7-point Likert scale. The uncertainty about mental states subscale (RFQ-u) captures poor use of mental state information and a stance characterized by a lack of knowledge about mental states. The certainty about mental states subscale (RFQ-c) captures better use of mental states information and adaptative levels of certainty about mental states. The French validation of the instrument demonstrated satisfactory reliability and construct validity of the two subscales [55].

<u>Experience in Close Relationships Scale-Revised (ECR-R)</u> [56]: a 36-item questionnaire of adult attachment style. The instrument, rated on a 7-point Likert scale, comprises two subscales measuring two attachment dimensions: anxiety (intense concern for relationships, fear of abandonment) and avoidance (feelings of discomfort in establishing emotional closeness with the partner, difficulty in trusting others). Four attachment styles can be defined according to individuals’ anxiety and avoidance scores [57,58]: “Secure” attachment style is associated with low scores on both anxiety and avoidance, “Fearful” is associated with high scores on both anxiety and avoidance, “Preoccupied” with high anxiety and low avoidance, and “Dismissing” with high avoidance and low anxiety. The French validation of the questionnaire has good reliability and validity and is consistent with its theoretical model of attachment dimensions [59,57,60].

<u>University of California Los Angeles Loneliness Scale [UCLA-LS)</u> [61]: a 20-item scale designed to measure one’s subjective feeling of loneliness and of social isolation on a 4-point Likert scale. Conceptualized as unidimensional in its structure, the scale items tap both the frequency and intensity of salient aspects and events of the lonely experience (e.g. How often do you feel alone ?). Evaluation of the scale found high internal consistency and good validity [62]. It is the most commonly used self-report loneliness instrument: a meta-analysis looking at 149 studies found that the UCLA scale was used in 27% of the studies, far more than for any other formal scale [62,63]. The French validation of the instrument demonstrated good internal consistency and construct validity [64].

<u>Symptom Checklist-90-Revised (SCL-90-R)</u> [65]: a 90-item instrument that evaluate a broad range of psychological problems and symptoms of psychopathology. On a 5-point Likert scale, it yields nine scores along primary symptom dimensions (somatization, obsessive-compulsive, interpersonal sensitivity, depression, anxiety, hostility, phobic anxiety, paranoid ideation, psychoticism) and three scores among global distress indices (global severity index - GSI, hardiness, symptom free). The French validation of the instrument demonstrated satisfactory reliability and validity of the French version of the SCL-90-R [66].

### Data analyses

We started the validation process of the French ETMCQ version with the translation of the original English ETMCQ by independent French and English Natives speakers (S1 File. ETMCQ items in French), following a forward-backward-forward procedure [67]. We conducted a Confirmatory Factor Analysis for the proposed three-factor model. The goodness-of-fit indices employed where: Chi-Square; the Comparative Fit Index (CFI), the Tucker-Lewis Index (TLI), the Standardized Root Mean Residual (SRMR), the Root Square Error of Approximation (RMSEA). To achieve a good fit of data, the values of CFI and TLI should be above .90 for an acceptable fit and above .95 for a good fit, the SRMR and the RMSEA should be under .08 for a, acceptable fit and under .05 for a good fit [68,69]. The internal coherence of the obtained subscales was estimated by calculating Cronbach’s alpha coefficients, which are deemed acceptable above .7. Intraclass correlation coefficients (ICC) were calculated for the 3 ETMCQ subscales to control for reliability. ICC above 0.74 was considered good, and between 0.60 and 0.74 acceptable [70]. Temporal stability was estimated using test-retest correlations after a one-year interval. Test-retest correlations above .7 were considered strong, above .6 good, and above .5 acceptable [71,72]. Construct validity was established by examining Spearman’s correlations between the ETMCQ subscales and 4 related constructs used in the original study [13]: childhood traumatic experiences (CTQ-SF), mentalization abilities (RFQ-8), attachment style (ECR-R). Additionally, correlation of the ETMCQ subscales with loneliness (UCLA-LS) and psychopathology (SCL-90-R) were also examined. Nonparametric Mann-Whitney U test for two independent groups evaluated difference in the ETMCQ subscale according to gender. One-way ANOVAs (Welch’s) and Dwass-Steel-Crichlow-Fligner pairwise comparisons were conducted to assess differences in ETMCQ subscales between the distinctive attachment styles as itemized by the ECR-R construct (“Secure”, “Fearful”, “Preoccupied” and “Dismissing”) [59], after a median split between of the ECR-R Avoidant and Anxious dimensions was done. A General Linear Model of Mediation evaluated the ETMCQ subscales mediation between childhood traumatic experiences (CTQ-SF total score) and psychopathology (SCL-90-R GSI), once considering first correlation between traumatic childhood experience and psychopathology. Another General Linear Model of Mediation evaluated the ETMCQ subscales mediation between loneliness (UCLA-LS) and psychopathology (SCL-90-R GSI), after considering correlation between loneliness and psychopathology. All p values were adjusted with Bonferroni corrections. All statistical analysis were performed using Jamovi Desktop 2.3.28.

## Results

### Demographic data

Our sample consists of 161 women (53.3 %) and 141 men (46.7 %). Mean age was 37.1 (standard deviation 12.3), with women mean age 37.3 (standard deviation 13.6) and men mean age 36.8 (standard deviation 10.7). Mean SCL-90-R GSI was .71 which is in line to what is found in general population [73–75]. Mean CTQ-SF total score was 42.5 corresponding to previous findings in non-clinical population [53,76].

### Factor structure, internal consistency and reliability

Table 1 gives the results of Confirmatory Factor Analysis (CFA). The initial 15-item 3-factor model did not provide good fit to the data (CFI .775; TLI .728; SMRR .0903; RMSEA .0984). Following the procedure of Campbell et al. [13], we removed in each ETMCQ subscale the item with the lowest loading (item 1 for EM: “I usually ask people for advice when I have a personal problem.”; item 3 for EM: “I’d prefer to find things out for myself on the internet rather than asking people for information.”; and item 6 for EC: “When I speak to different people, I find myself easily persuaded by what they say even if this is different from what I believed before.”). Goodness of fit was improved although not acceptable (CFI .815; TLI .760; SRMR.0666; RMSEA .0961). Next we correlated the residuals between several items, which gave satisfactory goodness of fit (CFI .954; TLI .935; SRMR .0480; RMSEA .0501). We allowed four Correlated Errors (CE) terms (or disturbances). While taking into account the problem of using CE in CFA framework [77], we based the choice of computing certain CE upon theoretical arguments (i.e. measurement model provided by the authors of the original instrument which includes CE terms with similar item content or wording: items 7&8, 5&12) and statistical ones (i.e. high modification indices and large standardized residuals in Jamovi: items 11&15, 13&14). Internal consistency was acceptable, with Cronbach’s alpha for the entire scale at .701, and for each ETMCQ subscale as follows: Trust .710; Mistrust .690; Credulity .742.

**Table 1.** Goodness-of-Fit measures.

| Model | CFI | TLI | SRMR | RMSEA |
| --- | --- | --- | --- | --- |
| 3-factor, 15 items | .775 | .728 | .0903 | .0984 (.09 - .10) |
| 3-factor, 12 items | .815 | .760 | .0666 | .0961 (.08 - .11) |
| 3-factor, 12 items, CR | .954 | .935 | .0480 | .0501 (.03 - .07) |

CRs = Correlated residuals; CFI = Comparative fit index; TLI = Tucker-Lewis Index; SRMR = Standardized Root Mean Square Residual; RMSEA = Root Mean Square Error of Approximation.

Mean ETMCQ subscale scores were the following: ET *M* = 4.81 (SD = 1.10); EM *M* = 3.76 (*SD* = 1.09); EC *M* = 2.54 (*SD* = 1.16). A nonparametric Mann-Whitney U test for two independent groups revealed that women and men differed on both ET and EC subscales, with women showing higher scores on both subscales (ET: women *M* = 4.79, *SD* = 1.08; men *M* = 4.45, *SD* = 1.1, *p* < .01; EC women *M* = 2.77, *SD* = 1.18; men *M* = 2.35, *SD* = .96, *p* < .01). Shapiro-Wilk test shown non-parametric distribution for ET (*p* < .001) and EC (*p* < 0.01). EM was close to a normal distribution (*p* = .05). When examining Spearman correlation coefficients between the ETMCQ subscales and age, we found a significantly negative correlation between age and EM (*r* = -.253, *p* < .001) and between age and ET (*r* = -.147, *p* < .05). All items load significantly (*p* < .001) and substantially (above .3) to their respective factors (Fig 1). EM and EC were significantly correlated (*r* = .339, *p* < .001), as were ET and EC (*r* = .170, *p* < .05). ET and EM were not significantly correlated (Fig 1).

**Fig 1.**
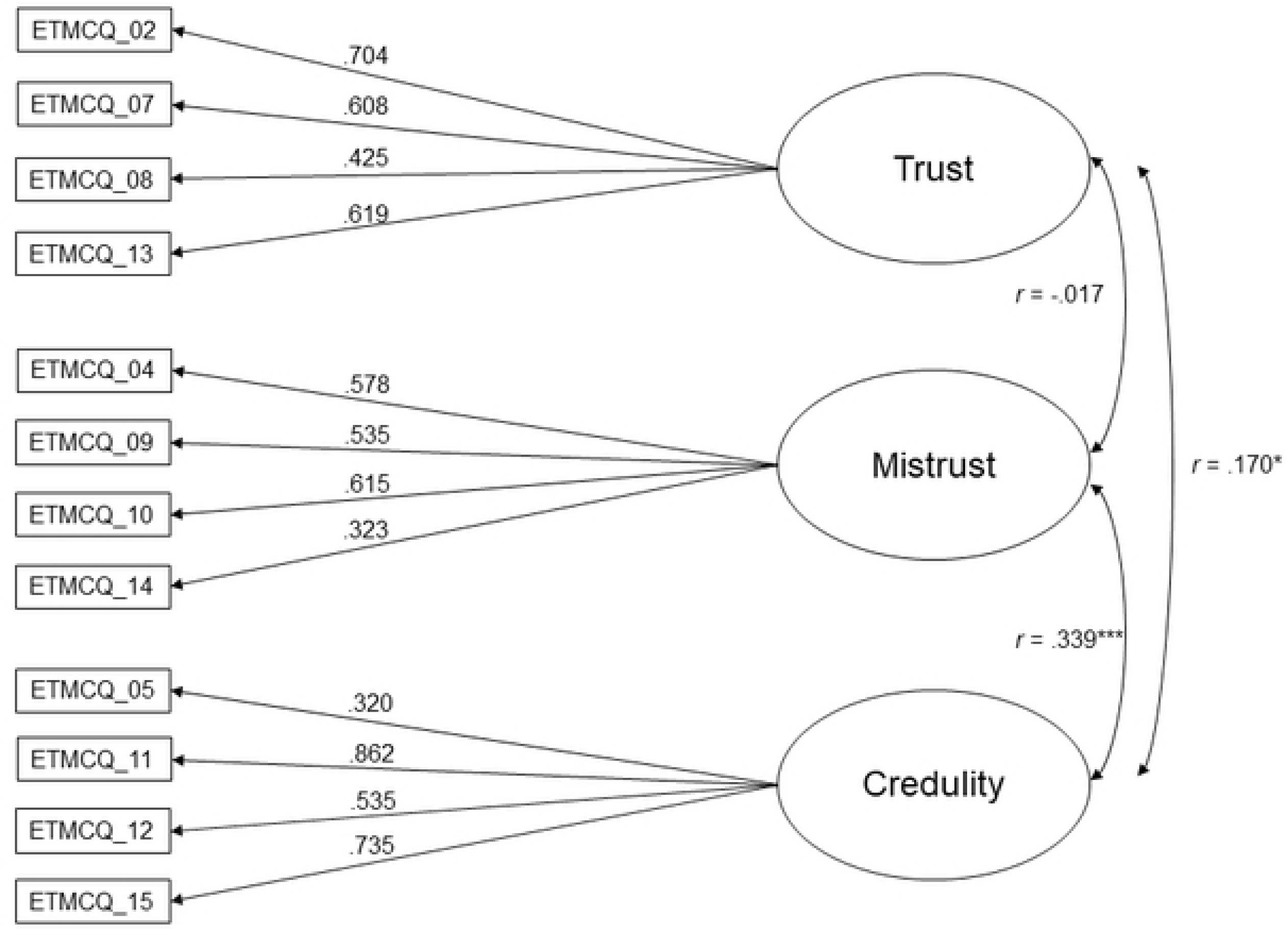
CFA with factor loadings (12 items). Loadings are standardized. Rectangles indicate measured variables and circles represent latent constructs. Note the item numbering is retained from the original Campbell study 15-item scale. * p < .05. ** p < .01. *** p < .001. All 302 participants were recontacted after a one-year interval, 257 took the retest (85.1% of the original sample). Between individuals who took the retest and those who did not, there was no significant differences in age (Mann-Whitney U = 5760.0, *p* = .97), gender (X^2^ (1) = 1.69, *p* = .19), GSI (Mann-Whitney U = 5261.0, *p* = .33) nor in the ETMCQ scores (ET Mann-Whitney U = 5079.5, *p* = .19, EM Mann-Whitney U = 5459.0, *p* = .55, EC Mann-Whitney U = 5144.0, *p* = .24). The test–retest reliability was moderate to good, with Spearman *rs* = .500 (*p* < .001), .600 (*p* < .001) and .678 for ET, EM and EC, respectively. Intraclass Correlation Coefficients (ICC) estimates and their 95% confident intervals were calculated based on average-rating (k = 257), absolute-agreement, 2-way random-effects mode [78,79]. ICC were acceptable to good: ET = .706 (95% CI (.62-.77)); EM = .733 (95% CI (58.-.82)) and EC = .781 (95% CI (.71-.83)).

### Construct validity

Significant correlations between the ETMCQ subscales and related developmental, psychological and psychopathology scales are given in Table 2.

**Table 2.** Spearman correlations between the ETMCQ subscales and convergent scales.

|  | ET | EM | EC |
| --- | --- | --- | --- |
| **Childhood traumas (CTQ-SF)** |  |  |  |
| Physical abuse | -.019 | .194* | .136 |
| Sexual abuse | .007 | .184* | .144 |
| Emotional abuse | .001 | .289*** | .305*** |
| Physical negligence | -.093 | .183* | .152 |
| Emotional negligence | -.123 | .270*** | .153 |
| Total score | -.067 | .300*** | .257*** |
| **Mentalization (RFQ-8)** |  |  |  |
| RFQ certitude | -.126 | -.324*** | -.496*** |
| RFQ incertitude | .161 | .280*** | .248*** |
| **Attachment (ECR-R)** |  |  |  |
| Avoidant dimension | -.342*** | .259*** | .104 |
| Anxious dimension | -.001 | .328*** | .430*** |
| **Loneliness (UCLA-LS)** |  |  |  |
| UCLA-LS sum | -.207** | .397*** | .231** |
| **Psychopathology (SCL-90-R)** |  |  |  |
| Global Severity Index | .118 | .396*** | .462*** |

P values adjusted for Bonferroni correction \* $p < .05$ . \*\* $p < .01$ . \*\*\* $p < .001$ .

Regarding childhood traumatic experiences as assessed in the CTQ-SF, EM was positively correlated with Physical Abuse (*r* = .194, *p* = < .05), Sexual Abuse (*r* = .184, *p* = < .05), Emotional Abuse (*r* = .289, *p* = < .001), Physical Negligence (*r* = .183, *p* = < .05) and Emotional Negligence (*r* = .270, *p* = < .001); EC was positively correlated with Emotional Abuse (*r* = .305, *p* = < .001); ET was not significantly correlated to any CTQ-SF subscale. EM was positively correlated to CTQ-SF total score (*r* = .300, *p* = < .001), as was EC (*r* = .257, *p* = < .001).

With regard to mentalizing abilities operationalized through the RFQ-8, EM was negatively correlated with the certitude subscale (*r* = -.324, *p* < .001) and positively with the incertitude subscale (*r* = .280, *p* < .001); EC was negatively correlated with the certitude subscale (*r* = - .496, *p* < .001) and positively with the incertitude subscale (*r* = .248, *p* < .001). ET was not significantly correlated to RFQ subscales.

Concerning attachment dimensions as reflected in the ECR-R scale, ET was negatively correlated with the avoidance dimension (*r* = -.342, *p* < .001); EM was positively correlated with the avoidance (*r* = .259, *p* < .001) and anxious dimension (*r* = .328, *p* < .001); EC was positively correlated with the anxious dimension (*r* = .430, *p* < .001).

With respect of psychopathology as itemized in the SCL-90-R instrument, EM was positively correlated with the GSI (*r* = .396, *p* < .001); EC was positively correlated with the GSI (*r* = .462, *p* < .001); ET was not significantly correlated with psychopathology. Note that when looking to the primary symptom dimensions, particularly strong and significant were the correlation between EM and paranoid ideation (*r* = .454, *p* < .10^-16^), as well as EC and interpersonal sensitivity (*r* = .464, *p* < .10^-16^), whereas ET was not significantly correlated with any of them. Regarding loneliness as operationalized in the UCLA-LS, loneliness score was negatively correlated with ET (*r* = -.207, *p* < .01); positively correlated with EM (*r* = .397, *p* < .001); and positively correlated with EC (*r* = .231, *p* < .01)

### ETMCQ and attachment

We assessed differences in the ETMCQ subscales between the distinctive attachment styles (“Secure”, “Fearful”, “Preoccupied” and “Dismissing”) [59]. A median split between of the ECR-R Avoidant and Anxious dimensions was done (median Avoidant = 74; median Anxious = 23), resulting in 101 participants being coded as having “Secure” attachment style (low avoidance and low anxiety), 103 “Fearful” (high avoidance, high anxiety), 51 “Preoccupied” (low avoidance, high anxiety), and 46 “Dismissing” (high avoidance, low anxiety). One-way ANOVAs (Welch’s) were conducted. ET subscale was found to differ significantly between the different attachment styles (ET F = 8.65, *p* < .001). Dwass-Steel-Crichlow-Fligner pairwise comparisons found that the “Secure” and “Preoccupied” styles scored significantly higher than the “Dismissing” and “Fearful” style (*ps* < .01). There were no significant differences between “Secure” and “Preoccupied” styles neither between “Dismissing” and “Fearful” style. EM subscale was also found to differ significantly between the different attachment styles (EM F = 12.19, *p* < .001). Pairwise comparisons showed significant difference between higher score for “Fearful” on both “Secure” (*p* < .001) and “Dismissive” styles (*p* < .05). Other comparisons did not significantly differ. Finally, EC subscale was found to differ significantly between the different attachment styles (EC F = 13.71, *p* < .001). Pairwise comparisons showed significant differences between higher score for “Fearful” and “Preoccupied” styles comparing to “Secure” and “Dismissive” ones (*ps* < .01). Other comparisons weren’t significantly different (Fig 2).

**Fig 2.**
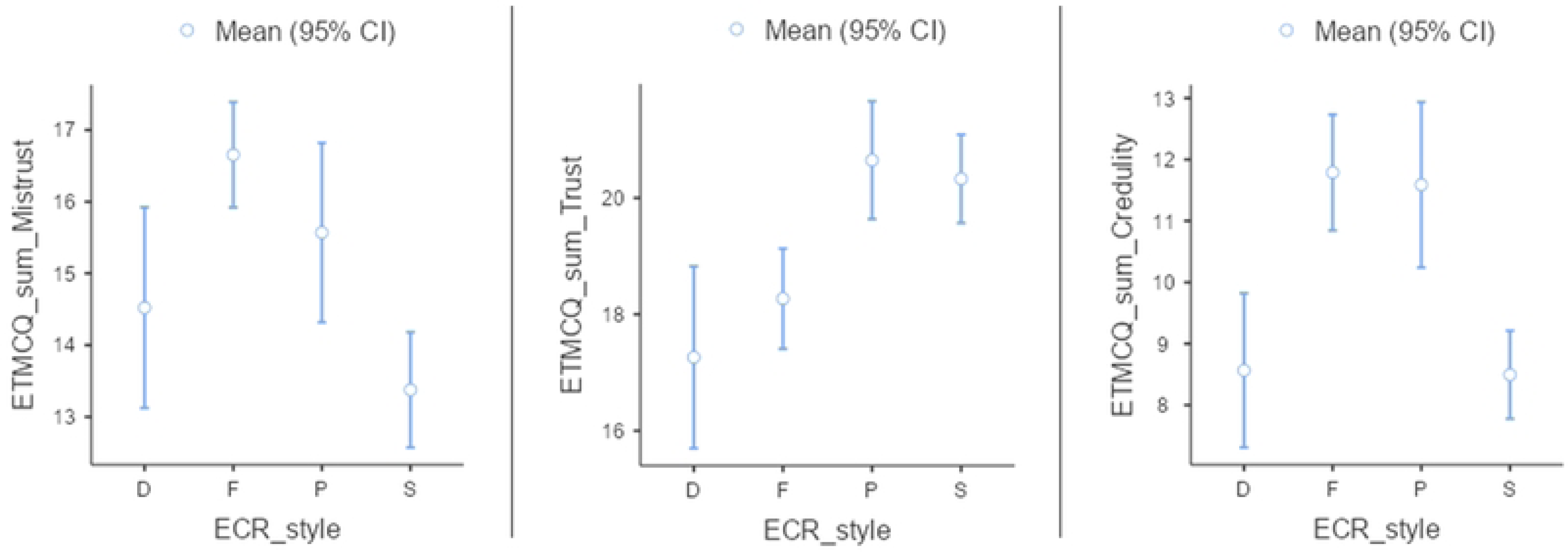
Comparison between to attachment styles according to the ETMCQ subscales. D: “Dismissive”. F: “Fearful”. P: “Preoccupied”. S: “ Secure”.

### ETMCQ and childhood traumatic experiences

A generalized linear model of mediation was constructed between CTQ-SF total score, the ETMCQ subscales and SCL-90-R GSI (Fig 3). We found that the total effect of CTQ-SF total score on GSI was significant (standardized effect sizes β = .278, *p* < .001). The direct effect of CTQ-SF total score on the GSI was significant (β = .133, *p* < .001), but smaller than the total effect, indicating partial mediation effects. As shown on the path diagram model of mediation (Fig 3), only EM and EC appear to mediate the relation between childhood traumatic experiences and psychopathology (EM standardized mediation effect between CTQ-SF total score and GSI : *β* = .066, *p* < .001 ; EC standardized mediation effect between CTQ-SF total score and GSI : *β* = .090, *p* < .001). ET had no significant mediating impact. It is to note that the when examining CTQ-SF subscales, Emotional abuse and Emotional negligence had the greatest direct effect on psychopathology (respectively *β* = .140, *p* < .005 and *β* = .121, *p* < .001), with EM and EC still keeping significant mediating effects.

**Fig 3.**
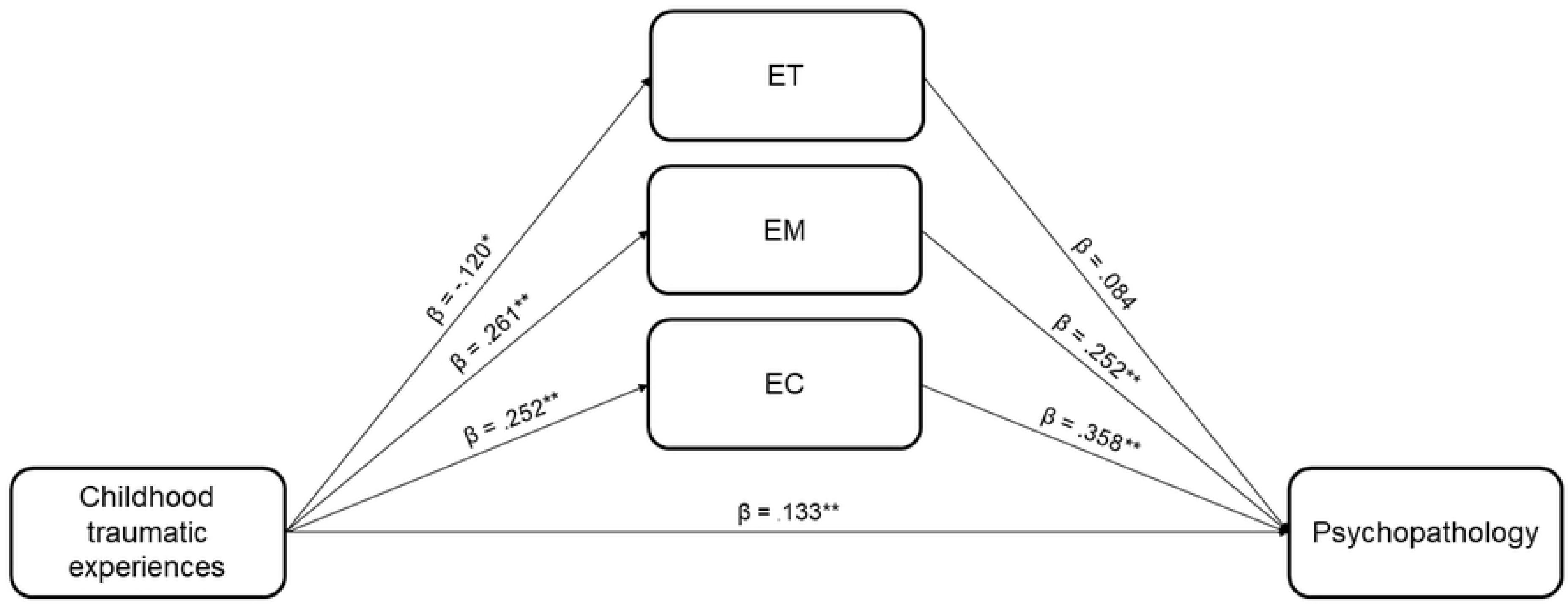
Path diagram model of mediation by the ETMCQ subscales on the relation between childhood traumatic experiences and psychopathology. β = standardized effect sizes * p < .05. ** p < .001

### ETMCQ and loneliness

A second generalized linear model of mediation was constructed between loneliness, the ETMCQ subscales and psychopathology (Fig 4). We found that the total effect of UCLA-LS sum on the SCL-90-R GSI was significant (standardized effect sizes β = .440, *p* < .001). The direct effect of UCLA-LS sum on the SCL-90-R GSI was significant (β = .337, *p* < .001), but smaller than the total effect, indicating partial mediation effects. Path diagram model of mediation by EM and EC on the relation between loneliness and psychopathology were significant and positive, and indicated a potential role of these factors as mediators of the relationship between loneliness and psychopathology (EM standardized mediation effect between UCLA-LS sum and SCL90-R GSI: *β* = .064, *p* < .01; EC standardized mediation effect between UCLA-LS sum and SCL90-R GSI: *β* = .076, *p* < .001). ET displayed a negative significant mediation effect (ET standardized mediation effect between UCLA-LS sum and SCL90-R GSI : *β* = -.036, *p* < .01) which indicate a likely suppressor effect, a situation in which the magnitude of the relationship between the independent variable and the dependent variable becomes larger when a mediating variable is introduced [80]. Thus, ET suppresses the relationship between loneliness and psychopathology when present and augments it when absent. Moderation tests were additionally conducted, indicating a significant effect for ET (*Z* = 2.096, *p*. <.05), while EM an EC did not moderate the association between loneliness and psychopathology.

**Fig 4.**
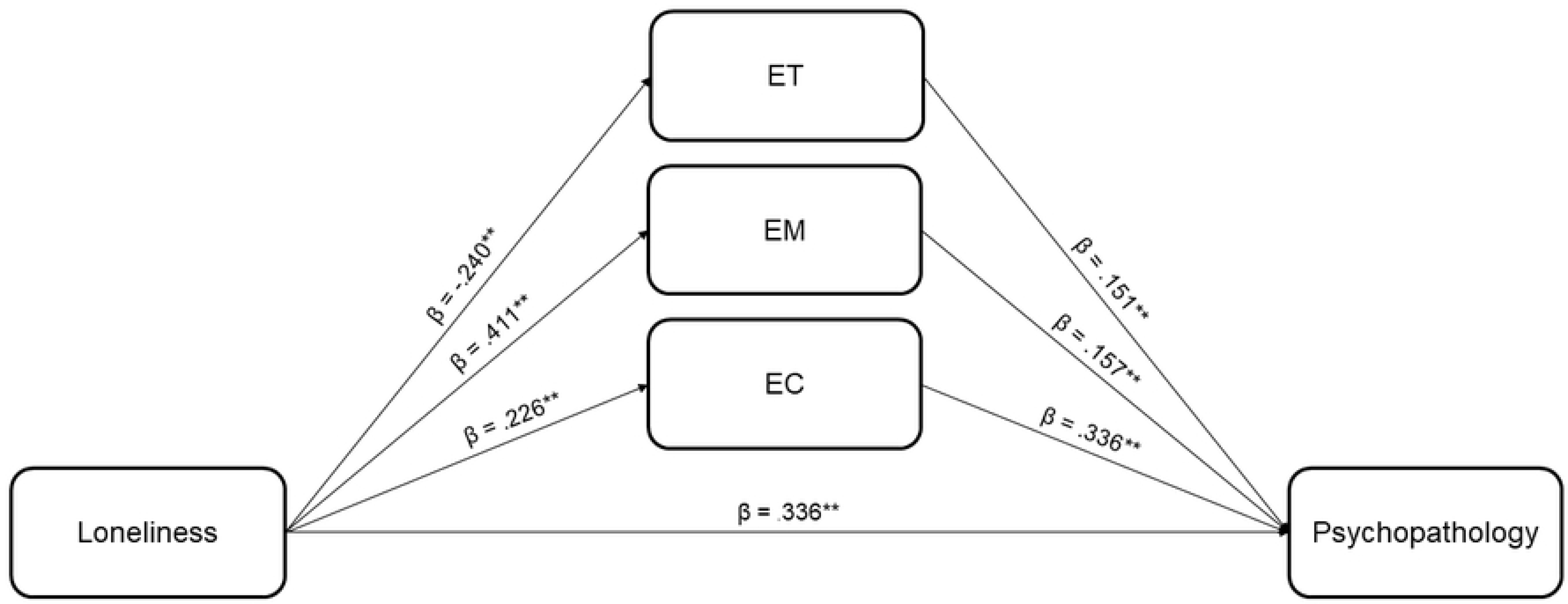
Path diagram model of mediation by the ETMCQ subscales on the relation between loneliness and psychopathology. β = standardized effect sizes ** p < .001.

## Discussion

Consistent with previous findings [19,13,18], the results of our study show that the ETMCQ represents a valid self-report instrument to assess epistemic trust. We find an adequate replication of the original three-factor solution in a francophone non-clinical population, with satisfactory psychometric properties and external validity. We further replicate on an independent sample Campbell et al. [13] findings on epistemic trust differences related to attachment, as well as the ETMCQ subscales mediating effect between childhood trauma experiences and psychopathology. We finally observe that ETMCQ subscales mediate the well-described association between loneliness and psychopathology.

Our analyses confirm a three-factor structure for a 12-item scale, referring respectively to the degree of trust (ET), mistrust (EM) and credulity (EC) in epistemic trust stances. To note that the three removed items present the lowest loadings in their respective subscales in the ETMCQ original and validation studies (item 1 for ET in Liotti et al. [19] and Asgarizadeh et al. [20], item 3 for EM in all three studies, item 6 for EC in Campbell et al. [13] and Liotti et al. [19]). We find significant associations among the three epistemic trust stances, with a slightly different correlation pattern than in the original Campbell et al. study [13]. We find gender differences between epistemic trust stances, with women showing higher scores on both ET and EC subscales. This result partially echoes Campbell et al. [13](who found also higher women EM score, but only in their Study 2), Benzi et al. [18] and Riedl et al. [27] (higher score only for ET), Asgarizadeh et al. [20](higher score for EC and EM), but not Liotti et al. [19] who found no differences between gender. We observe that age has an impact on epistemic trust stances, with ET and EM being negatively linked with age. This latter finding partially mirrors Campbell et al. [13] who find that ET (in their Study 1) and EC (in their Study 2) were negatively correlated with age. Liotti et al. [19] observed that only EM was negatively associated with age, and Asgarizadeh et al. [20] only ET. Internal consistency for the whole instrument and for the three subscales is acceptable, confirming findings from Campbell et al. [13] and Liotti et al. [19], but not Asgarizadeh et al. [20] who found low internal consistency for ET and EM. Test-retest reliability and interclass correlation coefficients of the subscales is acceptable to good, suggesting coherent and relatively stable individual attributes, reflecting possibly the mostly trait-driven nature of the ETMCQ measurement.

Concerning external validity, EM and EC subscales are significantly correlated with childhood traumatic experiences, insecure attachment, poor mentalizing, loneliness and psychopathology, adding weight to the hypothesis that they are both dysfunctional epistemic trust stances regarding social communication [13,81,82]. Furthermore, the fact that the three epistemic trust stances constitute independent factors was also supported by their distinctive correlation pattern with the precited variables. Interestingly, ET was not associated with reduced levels of symptoms nor most of the precited variables, except to avoidant attachment and loneliness. It adds weight to similar findings [23,18,26] that lead Li et al. [83] to formulate the hypothesis that higher level of the specific ET dimension is not correlated with better psychological functioning nor buffers against the impact of childhood adversity. Overall, we find mostly similar psychometric features with the original and the replicating ETMCQ validation studies, with possible differences in the number of items retained mainly due to linguistic and cultural factors that need further investigation.

On attachment issues, we find that the ETMCQ subscales differ according to distinctive attachment dimensions (avoidant and anxious). Our findings replicate those of Campbell et al. [13] and Asgarizadeh et al. [20] but not Liotti et al. [19], showing that EM is more strongly correlated to the avoidance dimension and EC to the anxiety dimension. In line with the novel way to see attachment as more adaptative than thought before, avoidance can be thought as adjustment to conditions promoting self-sufficiency, whereas anxiety may best aid in unpredictable environments [84]. Thus, avoidance may prompt EM and closing of the epistemic channel, whereas anxiety helps keep the channel open at the expense of self-agency [13,19]. We also replicate largely Campbell et al. [13] findings concerning attachment styles (“Secure”, “Fearful”, “Preoccupied”, “Dismissive”) and the ETMCQ subscales. “Secure” and “Preoccupied” styles scored higher than the other styles regarding ET, “Fearful” style higher regarding EM, and “Fearful” and “Preoccupied” higher regarding EC. Thus, “Dismissive” attachment style seems to protect against high level of EM and EC, but also show the lowest level of ET, hindering probably the possibility to learn from experience if a minimal level of the ET default mode is not attained [83]. “Fearful” attachment styles scores both high on EM and EC and low on ET, displaying the likely worst pattern of epistemic trust stances, eventually leading to the so-called “epistemic petrification” were all social-communicative channels are freezed [13,3]. “Preoccupied” attachment styles scores high on EM, EC and ET, thus having perhaps more likelihood to emerge from” epistemic petrification”.

Regarding childhood trauma experiences, we also replicate Campbell et al. [13] by observing that epistemic trust significantly mediates the association between childhood traumatic experiences and psychopathology. Specifically, we find that EM and EC partially mediate this link while ET was not a mediator, as observed by the authors of the original study. One possible interpretation could be that non-clinical sample are more protected against high levels of childhood traumatization, and thus further studies require looking into clinical populations to see if ET still lacks mediation effects. Preliminary studies report that ET is increased in clinical population, and it has been discussed that these findings may best be understood as indicating a help-seeking state rather than a trait [86]. When examining isolated childhood traumatic experiences, emotional abuse and emotional negligence had the greatest direct effect on psychopathology. Combined with our finding that EM and EC are mostly correlated to emotional abuse and negligence, they add evidence to the hypothesis that it is the emotional dimensions of child traumatization that has the most enduring impact on later life development, conceivably through its duration and enduring social-communicative disruptions [85,84,86]. Overall, our finding suggests that childhood traumatic experience, through adaptative attachment strategies (avoidance and anxiety dimensions), can produce dysfunctional epistemic stances (EM and EC) that participate to psychopathology vulnerability, possibly by hindering fundamental psychological abilities like mentalizing [17].

We further observe that the ETMCQ subscales significantly mediate the association between loneliness and psychopathology. Specifically, EM and EC are partial mediators, while ET has a suppressor effect, meaning that it has the potential to hinder paths between loneliness and psychopathology when present. These observations are corroborated by the fact that EM an EC did not moderate the association between loneliness and psychopathology, emphasizing thus the hypothesis that EM and EC have a primary mediating effect. The fact that ET shows this suppressor effect and is at the same negatively correlated to loneliness in our convergent validity testing specifies the shape of its peculiar participation in the epistemic stances frame. Thus, when ET is lacking, the person is presumably not able to open again to the social world - she/he is possibly protected against symptoms (no correlation between ET and psychopathology) but at the price of loneliness, stagnation, and the so-called “epistemic petrification”. Our results are in line with previous findings that showed that the development of epistemic trust capacities in childhood and adolescence is likely to be diminished in social contexts of loneliness, especially when there is social isolation from peers [87]. The phenomenon of “epistemic petrification” that occurs when epistemic channels are closed oblige to address the issues of enduring mistrust, suspicion and hopes that are always dashed that can submerge the thinking of individuals who experiences loneliness. Consequently, simply increasing individuals’ amount of social contact may be insufficient for improving outcomes. The development and dissemination of evidence-based interventions and primary prevention for loneliness is still in its infancy compared with recommendations for specific mental health disorders [88]. But what is known is that interventions that addressed maladaptive social cognition had a larger effect size compared to interventions that addressed social support, social skills, and opportunities for social contacts [89]. These results are consistent with both the Cacioppo at al. [47] model of loneliness as a self-reinforcing and regulatory loop, as with loneliness as an “epistemic petrification” precursor and marker. Some mental capacities are required in order for a person to see the value and engage in social relationships and in collective actions (so-called “we intentions”) [90,91]. In the social neuroscience field, underlying cognitive and biological processes have been found that could account for this bidirectional relation. These processes are possibly reduced for people undergoing prolonged loneliness, due to changes in biological features which can trigger fear-based responses to typically non-threatening situations, thus leading to loss of trust in relationships and further isolation [47,92,93]. Overall, this highlights the complex interaction between social support and mental health, individuals and communities, where social support could be a means to positive mental health, as well as the outcome of mental health improvement that can led to the capacity to engage further in social relationships [87].

Our study should be considered with its limitations in mind. First, our sample is community-based, and results and applications have to been experimented in clinical populations. Second, our study was operated through an online-survey. Although this method is becoming more widespread in the psychology research field, it may lead to the exclusion of part of the population, causing some limitations regarding the generalizability of our study. Third, our findings focused only on self-report measures and may introduce response biases potentially increasing the strength of reported associations. Future studies could assess the association between the ETMCQ and experimental designs which test laboratory social learning. Fourth, our study used a cross-sectional design and thus the direction of effect for the two mediation models cannot be formally establish. Accordingly, although the direction of effect in our first mediation model (epistemic trust mediating childhood traumatic experiences and psychopathology) is based on the assumption that childhood experience refers to the past while epistemic trust stances are current experiences, we cannot rule out alternative explanations such as selective recall of adverse events in individuals with current difficulties. In our second model (epistemic trust mediating loneliness and psychopathology), even if there is ample evidence from longitudinal studies that loneliness is a risk factor for multiple mental health outcomes [51], psychopathology produces also the experience of loneliness in itself. Therefore, as already mentioned by Campbell et al. [13], future longitudinal studies should test these directions and investigate the long-term effects of epistemic trust on mental health symptoms and psychological constructs.

## Conclusions

In line with the findings of Campbell et al. [13], Liotti et al. [19] and Asgarizadeh et al. [20], the results of our study show that the ETMCQ represents a valid and promising instrument to assess epistemic trust. We replicated Campbell et al. [13] main findings - which is valuable in itself considering the enduring replication crisis affecting psychology field [96] - concerning the ETMCQ subscales attachment style differences and mediating effect between childhood traumatic experiences and psychopathology. We lastly find that the ETMCQ subscales have a substantial mediating effect between loneliness and psychopathology. Thereby, we add momentum to the framework that considers epistemic trust as an underlying contributor to the maintenance or alleviation of psychopathology, through its freezing and in fine self-alienating effect on human development. Our study calls in favor of pursuing and enhancing research on the interceding role of epistemic trust in human difficulties. Future research should investigate epistemic trust in clinical population, where psychopathological expressions are severe, enduring and connected, and where identifying potential intercessors could help target and improve interventions [97].

## Data Availability

All relevant data are within the manuscript and its Supporting Information files.

## Supporting information

**S1 File. ETMCQ items in French.**

**S2 File. Study dataset 1.** ETMCQ french_for CFA

**S3 File. Study dataset 2.** ETMCQ french_for convergence

